# QVT Score, a radiomic biomarker of vascular complexity, enables prognostication and monitoring of NSCLC immunotherapy

**DOI:** 10.1101/2025.09.08.25335020

**Authors:** Young Kwae Chae, Vamsidhar Velcheti, Kai Zhang, Amogh Hiremath, Liam Il-Young Chung, Omid Haji-Maghsoudi, Rhea Chitalia, Jeeyeon Lee, Haojia Li, Seyoung Lee, Pushkar Mutha, Rushil Nagabhushan, David Levy, Diego Cantor, Yuchan Kim, Peter Haseok Kim, Amit Gupta, Trishan Arul, Anant Madabhushi, Nathaniel Braman

**Affiliations:** Feinberg School of Medicine, Northwestern University, Chicago, IL; Department of Hematology and Oncology, Mayo Clinic, Florida; Picture Health Inc, Cleveland, OH; Wallace H. Coulter Department of Biomedical Engineering, Georgia Institute of Technology and Emory University, Atlanta, GA, USA; Division of Cardiothoracic Imaging, Department of Radiology, University Hospitals Cleveland Medical Center, Cleveland, OH, USA; Department of Radiology, Case Western Reserve University School of Medicine, Cleveland, OH, USA

## Abstract

**Background:** Immune checkpoint inhibitors (ICIs) improve survival in advanced non-small cell lung cancer (NSCLC), yet current biomarkers such as PD-L1 expression and response criteria (RECIST v1.1) align poorly with long-term survival. Radiomics has been proposed as a source of novel biomarkers, but standard radiomic approaches suffer from limited biological interpretability and poor generalizability across treatment settings. We address these gaps by developing the Quantitative Vessel Tortuosity (QVT) Score, a biologically interpretable imaging biomarker that quantifies tumor vascular complexity -a known mediator of immune evasion - from routine imaging. We hypothesized that QVT Score would improve prognostication and enable treatment response monitoring in ICI-treated NSCLC, independent of current biomarkers.

**Methods:** This retrospective, multicenter study analyzed 1,301 CT scans from 682 ICI-treated NSCLC patients. An automated pipeline segmented lesions and tumor-associated vasculature within each scan, extracting 910 QVT features measuring vascular shape and complexity. Unsupervised clustering of these features in a discovery cohort (N=375) was performed to identify fundamental vascular phenotypes. A continuous QVT score was then derived using regularized logistic regression to map patients along this phenotypic spectrum. QVT Score was externally validated in ICI monotherapy (N=172) and chemoimmunotherapy (N=135) cohorts. In a longitudinal cohort (n=143), early on-treatment QVT Score changes were evaluated for overall survival (OS) association.

**Results:** Two robust vascular phenotypes emerged in the discovery cohort: a highly vascularized, chaotic “QVT High” phenotype with poor post-ICI OS and a “QVT Low” phenotype with normalized vasculature and improved ICI outcomes. The continuous QVT Score was prognostic for ICI monotherapy (HR = 1.17 per 0.1 increase, p = 0.0028) and chemoimmunotherapy (HR = 1.23 per 0.1 increase, p = 4.9×10⁻⁵). High QVT status remained prognostic for both treatments after adjustment for PD-L1 and clinical variables (adjusted HR range: 2.13–2.38, p ≤ 0.002). Early decreases in QVT Score during therapy, indicating vascular normalization, were associated with improved OS (HR = 1.93, p = 0.0022) independent of RECIST best overall response and tumor volume change.

**Conclusions:** QVT Score is a novel, biologically interpretable imaging biomarker that quantifies vascular complexity. It enables automated, non-invasive prediction and monitoring of ICI outcomes by capturing treatment-induced vascular remodeling. Integrating QVT Score into clinical decision-making and drug development can address critical gaps in precision oncology.

## Introduction

Immune checkpoint inhibitors (ICIs) have markedly transformed the therapeutic landscape of non-small cell lung cancer (NSCLC).^1–3^ However, a large portion of patients derive only limited or transient clinical benefit, underscoring a critical need for more robust predictive biomarkers.^4,5^ Existing biomarkers such as PD-L1, initially incorporated into clinical practice primarily to guide on-label treatment selection, have demonstrated suboptimal specificity and prognostic performance, leaving a substantial clinical gap in the stratification of patients for immunotherapy.^6^ In addition to this limitation in patient selection, there is also a dearth of effective means for monitoring treatment efficacy.^7^ Protocols for response monitoring on clinical imaging, such as RECIST evaluation, are well reported to have poor overall association to long term post-therapy outcomes, a misalignment that is especially pronounced in the immunotherapy setting.^8^

Disorganized tumor vasculature plays a critical role in fostering an immunosuppressive microenvironment by promoting hypoxia and impeding immune cell infiltration, which directly limit the efficacy of ICIs.^9^ Quantitative assessments of vascular complexity are crucial for elucidating primary resistance mechanisms to ICIs. Notably, impaired vascular architecture not only predicts ICI resistance, but also serves as a potential criterion for identifying patients who might benefit from therapeutic escalation, including the addition of anti-angiogenic agents to restore immune accessibility.^10^ The lack of robust and non-invasive tools to characterize tumor vasculature hinders both the prediction of ICI responses and precise patient stratification for combination therapies.^11^

Radiomics, the high-throughput extraction of quantitative imaging features from medical images, has emerged as a powerful approach for precision oncology biomarkers.^12^ A number of studies have explored such techniques for ICI prediction and monitoring, but barriers to clinical adoption persist.^13^ Many radiomic studies to date have focused on highly customized predictive algorithms designed for specific clinical indications. While these approaches can yield strong performance, they often lack the broader clinical utility of biomarkers targeting well-characterized biological mechanisms, such as gene panels or immunohistochemistry assays. Additionally, traditional radiomic features also frequently rely on techniques developed for general computer vision and do not account for biological context, limiting interpretability and translational impact.^14,15^ Deep learning-based models have been applied for precision oncology but such approaches further exacerbate interpretability challenges, as their predictions depend on complex, abstract feature representations with no connection to known disease biology.^16,17^ Accordingly, such deep learning models cannot provide mechanistic insight into treatment outcomes and are often prone to concealed biases.^18^

Quantitative Vessel Tortuosity (QVT) is a class of radiomics features developed to characterize tumor vascular complexity on standard CT imaging, which measures patterns of abnormal vascular growth such as vessel twisting and erratic branching. QVT has been shown to be associated with malignancy risk, adverse events, and responses to a variety of treatment strategies in multiple cancers and imaging modalities.^19–21^ In the ICI setting, Alilou et al. demonstrated associations between both the baseline values and on-treatment changes of individual QVT to immunotherapy outcomes in NSCLC, suggesting a direct link between imaging features and biologically driven treatment response mechanisms.^19^ However, prior QVT studies relied on individual feature associations or indication-specific models, limiting clinical applicability and failing to provide a single, biologically interpretable score generalizable across treatment contexts and imaging timepoints.^22^

Building upon this foundation, we sought to establish and validate (Figure 1) a more comprehensive and clinically generalizable radiomic biomarker: one that summarizes the QVT feature class to capture intrinsic vascular phenotypes and enables continuous scoring for both personalized prediction and monitoring. Unlike prior QVT research,^19–21,23,24^ which explored radiomic models customized for each new indication, we hypothesized that a biomarker directly characterizing the overall vascular landscape of NSCLC would have broader association with ICI outcomes. Through unsupervised clustering of hundreds of hundreds of individual interpretable radiomic QVT features, we identified two reproducible, intrinsic vascular phenotypes in NSCLC - one characterized by simpler normalized vascularity, and the other by complex, highly tortuous, chaotic vascular architecture - with associations to ICI outcomes. From these, we developed a continuous biomarker, QVT Score, to quantify tumor vascularity across a spectrum spanning these two distinct biological states. Furthermore, we developed a fully automated, cloud-based radiomic biomarker pipeline, enabling QVT Score to be seamlessly calculated from standard clinical imaging and applied for both single timepoint prognostication and longitudinal monitoring of treatment-induced vascular changes. In this paper, we evaluate this tool for association with survival to multiple ICI-based therapeutic regimens and its adjunctive value relative to established biomarkers and endpoints.

**Figure 1.**
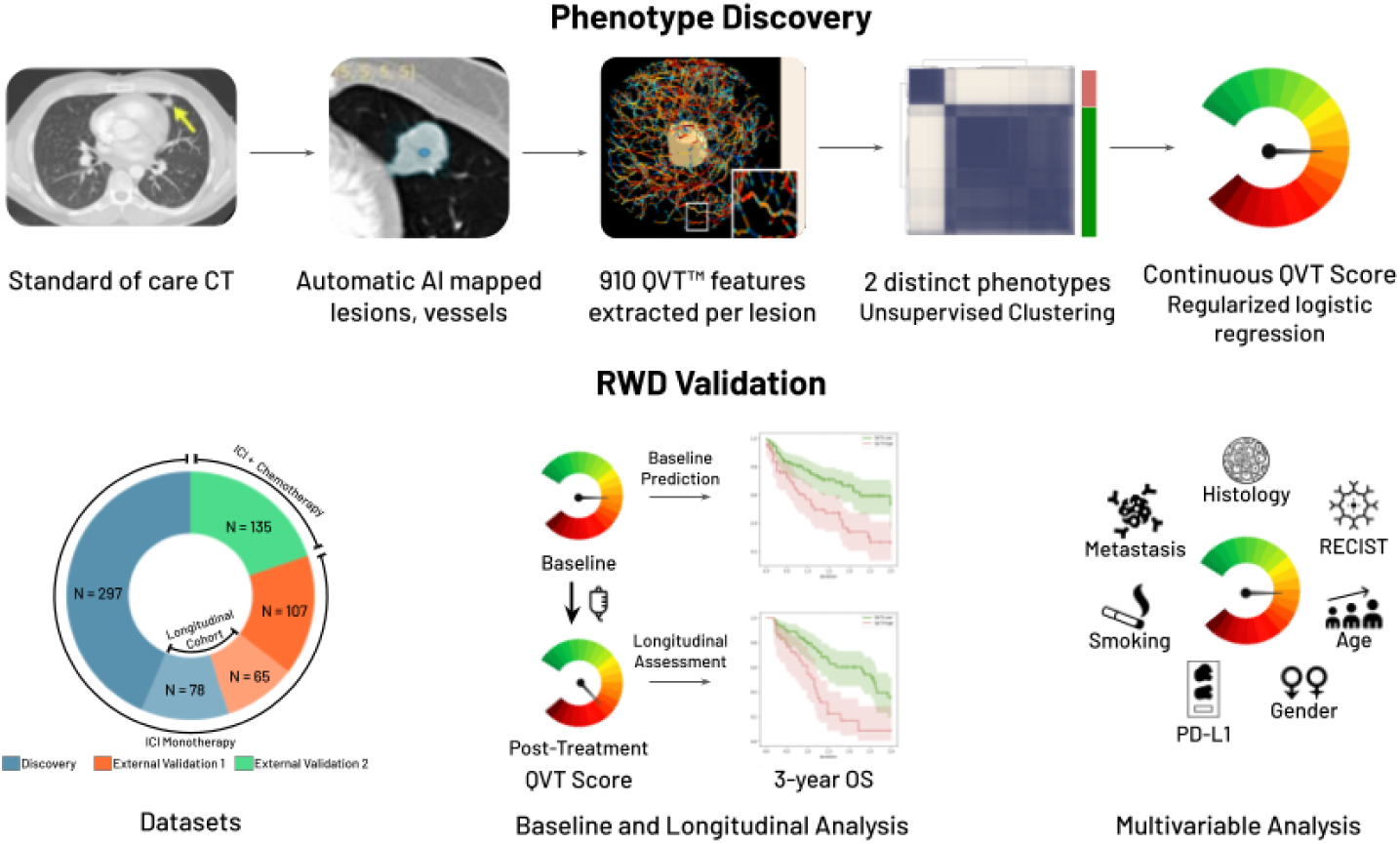
QVT™ Phenotype Discovery and Validation Workflow. Top: phenotype discovery procedures, bottom: real world data validation and analyses.

## Materials and Methods

### Study Population

An internal real-world database consisting of clinical data and radiology studies was queried for advanced NSCLC patients who received an ICI alone or in combination, returned a total of 7,563 image series acquired during 2,048 unique image studies of 854 unique patients for potential eligibility. The dataset featured data from 6 institutions, and was comprised, in part, of data published in prior radiomics studies.^19,23–25^

Inclusion criteria included the availability of a pretreatment and/or first on-treatment imaging study, one or more chest CT scans with visible presence of pulmonary lesions, and the availability of overall survival data. Exclusion criteria included patients who received radiation alongside ICIs, patients with uncertain treatment timing relative to imaging, and imaging data lacking critical metadata (e.g. pixel spacing and slice thickness).

This resulted in a final cohort of 1,301 CT image series collected during 825 imaging studies from 682 patients with advanced NSCLC treated who received treatment with ICIs, given with or without chemotherapy. All imaging and clinical data was collected under IRB-approved protocols with appropriate patient consent or waiver. Clinicodemographic details are summarized in Table 1.

**Table 1.** Patient demographics and clinical characteristics for discovery and validation cohorts.

| Characteristic |  | Discovery Cohort | External Validation Cohorts |  | Longitudinal Validation Cohort |
| --- | --- | --- | --- | --- | --- |
|  |  | ICI Monotherapy (n=375) | ICI Monotherapy (n=172) | Chemo-ICI (n=135) | ICI Monotherapy (n=143) |
| Age, mean (SD) |  | 66.4 (10.2) | 69.2 (11.5) | 66.1 (10.7) | 64.6 (11.3) |
| Sex, n (%) | Male | 176 (46.9) | 86 (50.0) | 49 (36.3) | 71 (49.7) |
|  | Female | 192 (51.2) | 86 (50.0) | 67 (49.6) | 72 (50.3) |
|  | Unknown | 7 (1.9) | 0 (0.0) | 19 (14.1) | 0 (0.0) |
| Histology, n (%) | Adenocarcinoma | 245 (65.3) | 117 (68.0) | 90 (66.7) | 95 (66.4) |
|  | Squamous Cell Carcinoma | 67 (17.9) | 21 (12.2) | 11 (8.1) | 20 (14.0) |
|  | Other | 61 (16.3) | 34 (19.8) | 15 (11.1) | 28 (19.6) |
|  | Unknown | 2 (0.5) | 0 (0.0) | 19 (14.1) | 0 (0.0) |
| PDL1 expression, n (%) | High | 67 (17.9) | 61 (35.5) | 19 (14.1) | 12 (8.4) |
|  | Positive | 32 (8.5) | 24 (14.0) | 43 (31.9) | 21 (14.7) |
|  | Negative | 86 (22.9) | 22 (12.8) | 42 (31.1) | 14 (9.8) |
|  | Unknown | 190 (50.7) | 65 (37.8) | 31 (23.0) | 96 (67.1) |
| Metastasis, n (%) | Yes | 323 (86.1) | 139 (80.8) | 105 (77.8) | 112 (78.3) |
|  | No | 52 (13.9) | 33 (19.2) | 30 (22.2) | 31 (21.7) |
| Smoking history, n (%) | Current | 86 (22.9) | 12 (7.0) | 15 (11.1) | 20 (14.0) |
|  | Past | 231 (61.6) | 130 (75.6) | 79 (58.5) | 98 (68.5) |
|  | Non-Smoker | 51 (13.6) | 30 (17.4) | 22 (16.3) | 25 (17.5) |
|  | Unknown | 7 (1.9) | 0 (0.0) | 19 (14.1) | 0 (0.0) |

#### Discovery Cohort

A discovery cohort of 375 patients was defined as the set of all patients treated at Institution A (n=226), Institution B (n=87), or Institution C (n=62). All patients in the Discovery cohort received ICI monotherapy.

#### External Baseline Validation Cohorts

Two external validation cohorts were used to independently validate the QVT Score in multiple ICI-based therapeutic strategies using imaging data from unseen institutions. These datasets were used to separately evaluate the association of QVT Score with outcomes of ICI monotherapy and chemoimmunotherapy.

- **ICI Monotherapy Validation Cohort:** Included 172 NSCLC patients (Institution D: n=96, Institution E: n=76) treated with ICI monotherapy from two additional institutions.
- **Chemo-ICI Validation Cohort:** Included 135 NSCLC patients (Institution D: n=58, Institution E: n=58, Institution F: n=19) who received combination chemo-immunotherapy.

#### Longitudinal Cohort

A longitudinal subcohort was constructed from the set of all ICI-treated patients with longitudinal CT data also available, consisting of 143 patients who had both pre-treatment and early on-treatment CT scans. This cohort combined 78 patients (Institution B: n=73, Institution C: n=5) from the Discovery cohort and 65 patients from the Validation cohort (Institution E: n=65). All patients received ICI monotherapy. This cohort was used to evaluate temporal changes in vascular features and QVT Score dynamics.

### Image Annotation and Preprocessing

#### Preprocessing and standardization

All CT images were resampled to a standard axial spacing of 0.7 x 0.7 mm and slice thickness of 1.25 mm to ensure spatial consistency in the downstream analysis. A full body organ segmentation model was applied to annotate the lungs and crop the image to their boundaries.

#### Automated lesion and vessel segmentation

Lesion segmentation was performed using a dedicated deep learning model trained to identify and delineate pulmonary lesions. Pulmonary vessels were further segmented using a separate automated vessel segmentation model. All segmentation output masks were visually assessed by experts for quality assurance; segmentations flagged for inaccuracy were corrected or excluded from further analysis. A total of 2,211 lesions were identified and delineated using this pipeline.

### QVT Feature Extraction

For each lesion, a peritumoral region was centered on the lesion for analysis of surrounding tumor-associated vasculature. The tumor vasculature was quantified by creating a skeletonized representation of the segmented vessel network. This is done by identifying the centerlines of each vessel and dividing the vascular network into branches. A comprehensive set of 910 QVT features were extracted, encompassing morphological parameters such as curvature along vascular branches, analysis of branching patterns, as well as characterization of vessel dimensional and volumetric attributes. For patients with more than one lesion or image series, lesion-level QVT features were averaged to obtain a single patient level set of summarized QVT features.

### Phenotype Discovery and Scoring

#### Unsupervised Clustering

QVT features from pre-treatment CTs from the discovery cohort were subjected to hierarchical consensus clustering^26^ for 250 iterations with 75% random sampling without replacement. The optimal number of clusters (k) was identified based on area under the curve (AUC) of the cumulative distribution function (CDF).

To derive final cluster assignments for the discovery cohort, K-means clustering was applied to the consensus matrix.

#### Development of the QVT Score

To enable more granular characterization of tumor vascular complexity, we derived a QVT Score by training a regularized logistic regression classifier to predict vascular phenotype within the discovery cohort using QVT features.

This process created the QVT Score, an interpretable radiomic biomarker of vascular complexity for prediction and monitoring of ICI-based treatments. The score summarizes a total of 910 QVT features into a single continuous measure (0–1) of vascular complexity, with higher scores more closely corresponding to a phenotype of high vascular complexity.

Thresholds for stratifying QVT Score into the corresponding vascular phenotypes were set within the discovery cohort in order to maximize the accuracy of phenotype classification.

#### Cloud-based platform for QVT Scoring

A fully automated QVT scoring pipeline was developed and deployed within a cloud-based architecture on Amazon Web Services (AWS). This end-to-end pipeline integrates organ segmentation, lesion segmentation, vessel segmentation, QVT feature extraction, and QVT score computation. Task orchestration and monitoring were managed using ClearML, enabling scalable and reproducible execution. To assess pipeline efficiency, we measured total processing time from raw image input to final QVT score output on CT data from a random sample of 60 patients.

### Longitudinal Analysis

For patients in the longitudinal cohort, both pretreatment and first on-treatment scans were processed using the QVT radiomics pipeline and QVT Score was computed for each timepoint. The change in QVT Score was computed as the difference between the on-treatment and baseline scores to quantify the extent of on-treatment change in vascularity.

The change in QVT Scores was thresholded by the median value within the longitudinal set to derive two QVT change groups, corresponding to patients whose vascularity increased following treatment (QVT Increasing) and those whose vascularity was reduced by immunotherapy (QVT Decreasing).

### Statistical Analysis

QVT Scores (baseline QVT Score, followup QVT Score, change in QVT Score) and QVT-derived patient groups (baseline QVT High/Low status, followup QVT High/Low status, and QVT Score Increasing vs. Decreasing groups), were evaluated for association with OS. Kaplan–Meier curves, log-rank tests, and multivariable Cox proportional hazards models were used to evaluate the association of QVT phenotypes and scores with survival outcomes. The primary metric of OS association was hazard ratio (HR), along with associated confidence intervals (CIs). Significance testing was performed according to the log rank test. Due to heterogeneity in the duration of followup across the multi-institutional real world datasets, survival analysis was restricted to 3-year OS.

To estimate its adjunctive benefit with respect to other predictive baseline variables and longitudinal endpoints, we examined QVT Score’s adjusted HRs and p values when included within a multivariable Cox Proportional Hazards model. Baseline QVT Score was compared with age, sex, PD-L1 tumor proportion score (TPS), presence of metastasis, and smoking history. Delta QVT Score was compared against tumor volume and RECIST response definitions (Objective Response [PD or SD vs. PR or CR], Disease control [[PD vs. SD, PR, or CR]). To handle missingness within clinical variables, we performed multiple imputation with chained equations and pooled hazard ratios and p-values across 30 iterations according to Rubin’s Rule.^27,28^

## Results

### Intrinsic Vascular Phenotypes in NSCLC

Unsupervised clustering of all QVT features in the Discovery cohort revealed distinct vasculature clusters, interpreted as phenotypes. Optimal clustering, as measured by AUC of the CDF, was achieved with two clusters (k=2). A minority cluster, containing 21.6% of patients, was characterized by elevated expression of QVT features corresponding to complex vascularity, and was accordingly denoted as “QVT High”. The other 78.4% of patients fell into the “QVT Low” cluster, marked by more ordered and normalized vasculature relative to QVT High patients. “QVT Low” was distinguished by reduced expression of QVT High-associated features, as well as elevated values of features associated with a more ordered vascular network. Representative examples of QVT feature expression for these groups are visualized in Figure 2. These vascular phenotypes were strongly associated with long term outcomes. The QVT High group was associated with significantly worse OS (HR = 1.65, p = 0.04).

**Figure 2.**
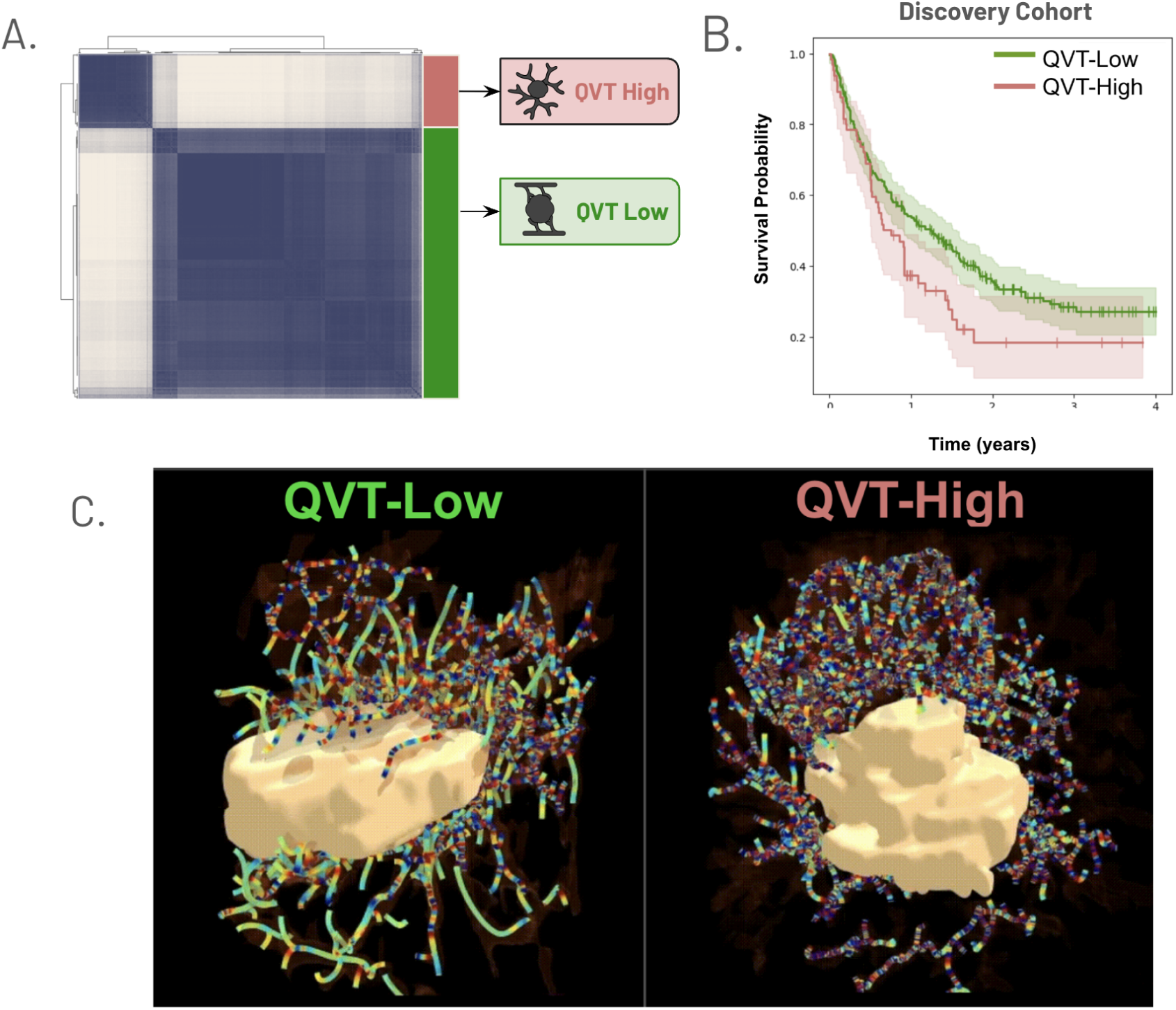
Characterizing intrinsic vascular phenotypes from QVT radiomics. a) unsupervised hierarchical consensus clustering of 910 radiomic QVT features reveals two distinct clusters. b) Kaplan Meier curves reveal baseline vascular phenotypes associated with outcomes on ICIs. c) Representative visualization of a QVT feature associated with vascular complexity, one of the individual features contributing to QVT Score. Patients with low expression of this feature have reduced QVT Score and more favorable OS. Patients with elevated QVT Score, have increased expression of features related to abnormal vessel shape at baseline - indicating vascular complexity - and greater hazard of death.

### QVT Score: A Radiomic Vascular Biomarker

The QVT score was developed to provide a single-variable, granular characterization of tumor vascular complexity, expanding on the innate biology discovered through vascular phenotyping. A threshold of 0.70 on QVT scores most accurately delineated the original vascular phenotypes in the discovery cohort. This threshold correctly identified QVT phenotype within the discovery with an AUC of 0.998 and an accuracy of 0.989. The average time required for fully automated QVT Scoring was 28.6 minutes (range: 15.6 minutes to 73.3 minutes).

The features with the strongest contribution to QVT Score were investigated based on the coefficients of the linear regression model. Features related to branching patterns, vessel curvature, and vessel twisting, were among the features associated with elevated QVT Score.

**Figure 3.**
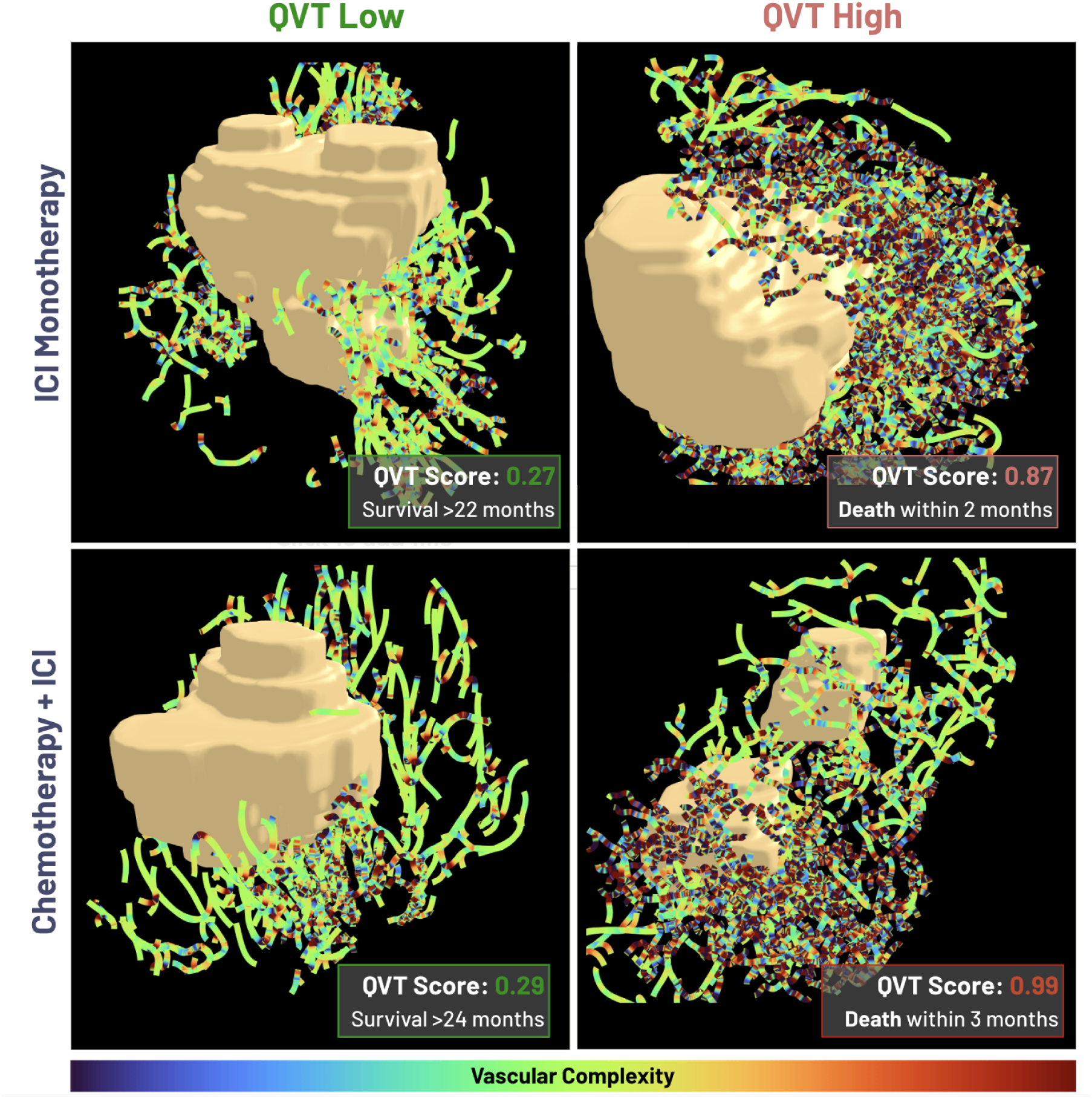
Visualization of curvature gradient feature in representative patients correctly stratified by QVT Score groups, but not PD-L1 status. Visualizations depict QVT curvature gradient, a vascular complexity-associated feature expressed in QVT Score High patients.

### QVT Score predicts outcomes for multiple immunotherapy regimens at baseline in external validation

QVT Score was evaluated to confirm its ability to predict outcomes to multiple ICI-based regimens in NSCLC using external testing datasets. As in the discovery cohort, both the continuous QVT Score (HR=1.17 per 0.1 increase, p=0.0028) and the QVT Score High group (HR=2.07 p=0.0022) were significantly predictive in the ICI Monotherapy testing cohort. Median OS was substantially reduced in the QVT Score High group (14.0 months), as compared to QVT Score Low, where the median OS was not reached due to insufficient death events at 3 years. Furthermore, QVT Score was also highly associated with OS in the Chemoimmunotherapy testing cohort with HR=1.23 per 0.1 score increase (p=4.9e-05). The QVT Score High group was associated with poorer post-combination therapy survival (HR=2.77, p=7.7e-05), as well as reduced OS (13.9 months).as compared to QVT Score Low (31.6 months). QVT Score was also associated with OS for the combined all comers testing cohort with HR=1.18 per 0.1 score increase (p<1e-05) and HR=2.26 (p<1e-5) for the QVT Score high group. Median OS among all-comers was 14.0 and 35.1 months for the QVT Score High and Low groups, respectively. Kaplan Meier curves depicting the OS stratification are included in Figure 4. In a subset analysis exploring prognostication in PD-L1 expression groups, QVT Score remained significant for PD-L1 High ICI Monotherapy recipients and for PD-L1 positive and negative chemoimmunotherapy recipients (Supplementary Figure 1).

**Figure 4.**
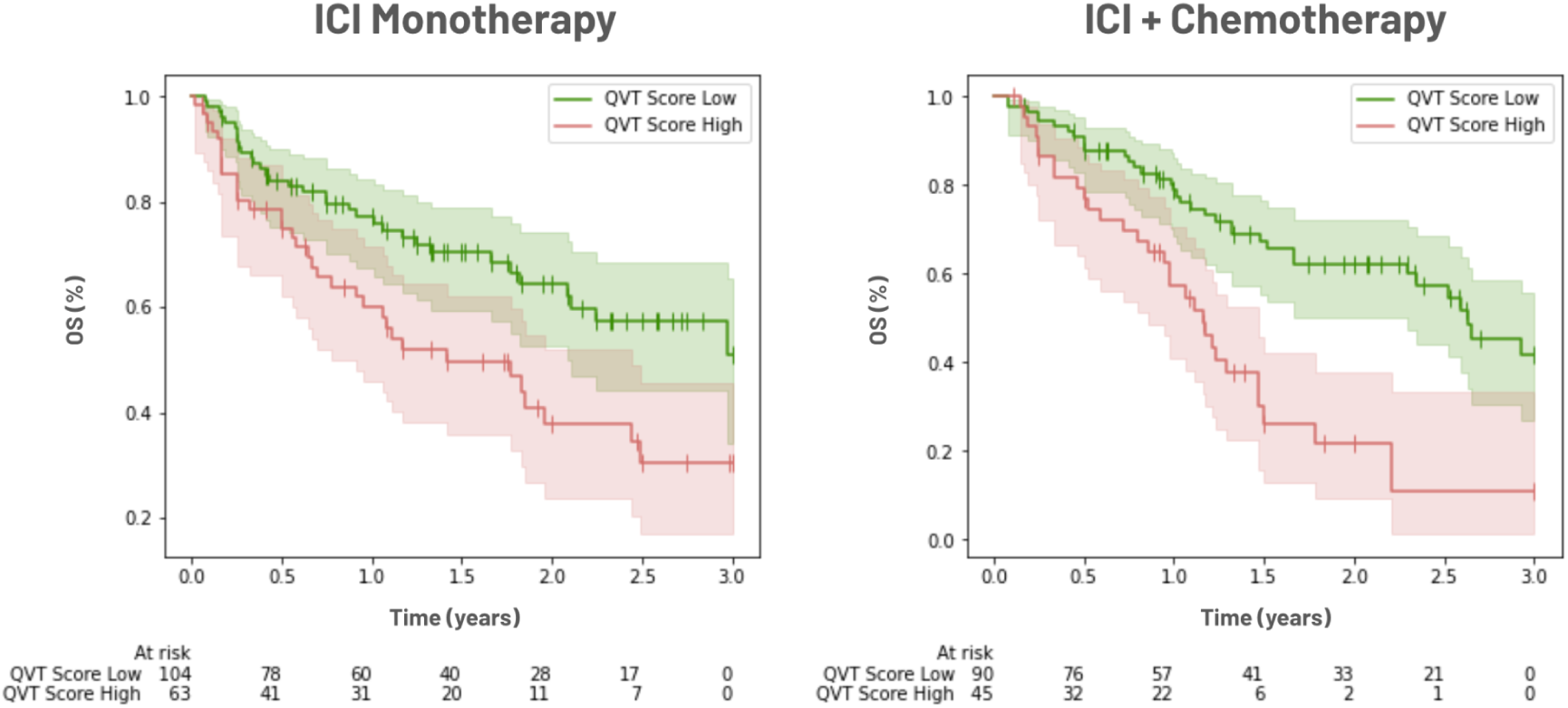
Kaplan Meier curves comparing OS stratification of baseline QVT Score categories for multiple ICI-based regimens in the external testing sets.

To evaluate the QVT Score’s ability to serve as an adjunctive biomarker for NSCLC treatment guidance, we performed a multivariable comparison for OS association with other baseline variables using a Cox Proportional Hazards model. While some clinical variables, including PD-L1 status and smoking history, were OS-associated in some cohorts, only QVT Score groups remained predictive of outcomes for all treatment groups. QVT Score High remained OS-associated independent of baseline variables, with adjusted HRs of 2.13 (p=0.0019) for ICI monotherapy, 2.38 (p=0.001) for combination therapy, and 2.28 (p<1e-5) for the all ICI treatments combined.

**Figure 5.**
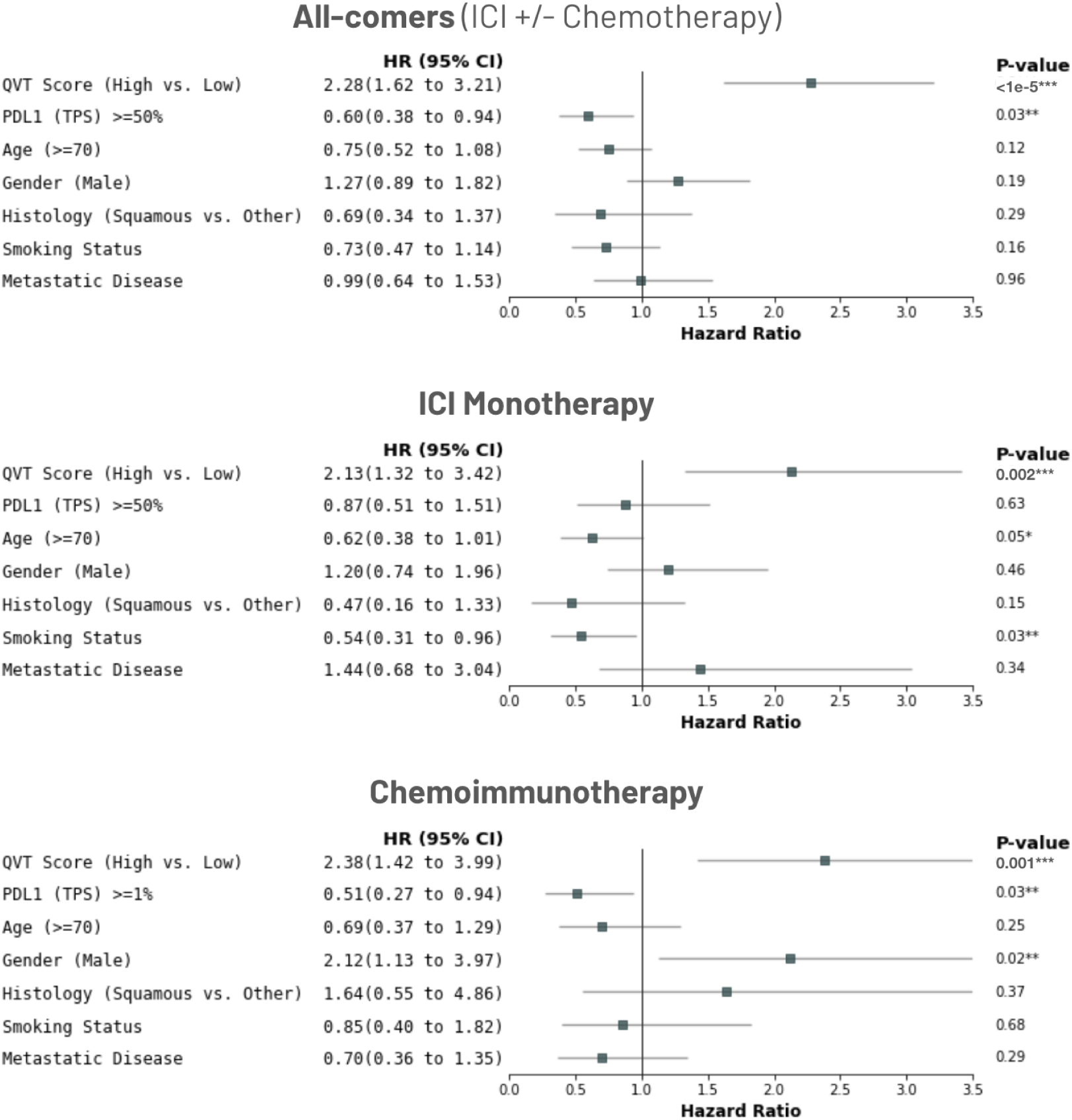
Forest plots showing association of the QVT Score high group with OS in the external testing sets following multivariable adjustment for baseline clinical variables. Multiple imputation with chained equations was used to account for variables with missing data. For the treatment-specific test sets, PD-L1 threshold was chosen according to the most established cutoff for that indication.

### Longitudinal change in QVT Score predicts survival benefit independent of size-based response criteria

To capture treatment-induced changes, we computed the difference between the on-treatment and baseline QVT Scores (Figure 6). A positive change in QVT Score was significantly associated with worse OS (HR=1.11 per 0.1 increase, p=0.012). Using the median change in QVT Score (0.0297) as a threshold, patients classified as QVT Score Increasing showed significantly poorer OS compared to those with decreasing scores (HR=1.93, p=0.0022), indicating an association between score reduction and vascular normalization. Median OS was 22.0 months for patients with decreasing QVT Score, as compared to a median OS of 9.0 months for the QVT Increasing group.

**Figure 6.**
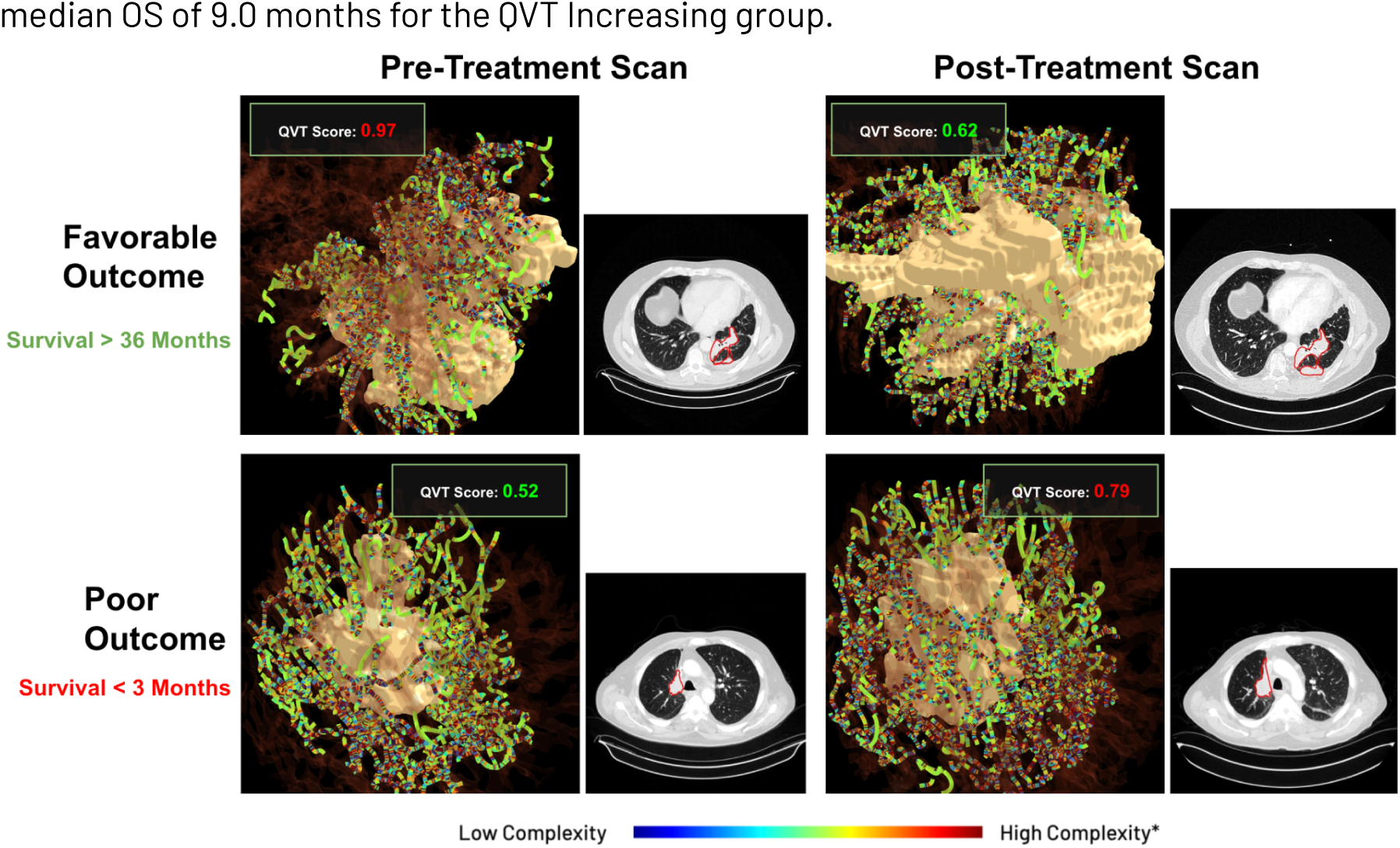
Treatment-induced vascular changes on first followup imaging are associated with survival following ICI monotherapy, as measured by Delta QVT Score. Visualizations of a feature emphasizing vessel twisting show an on-treatment decrease of vascular complexity is associated with favorable OS, while increasing vascular complexity indicates mortality risk.

We next evaluated whether the change in QVT Score was adjunctive to established size-based response criteria (RECIST v1.1 best overall response) and tumor volume. Following multivariable comparison, adjusted HRs for QVT Score groups remained significantly prognostic with respect to objective response (OR) (HR=1.67, p=0.018), disease control (HR=1.55, p=0.044) and tumor volume change (HR=1.84, p=0.0044),. Both tumor volume (Decrease vs. Increase: HR=1.64, p=0.018) and RECIST response (OR: HR=0.35, p=1e-4; Disease Control: HR=0.28, p<1e-5) were also significant in the multivariable comparison.

To further determine the clinical value of QVT dynamics, we evaluated whether longitudinal monitoring of QVT Score change provided additional prognostic value beyond a baseline QVT Score assessment. Both the baseline QVT Score (Adjusted HR = 2.27, p = 0.0027) and treatment induced QVT Score change (HR = 2.65, p = 0.00009) were independently associated with OS in a multivariable comparison. Furthermore, the prognostic impact of QVT Score change was magnified when patients were stratified by baseline QVT subgroups. Among patients who were QVT Low at baseline (n=109), those who later had an on-treatment increase in QVT Score were at substantially higher risk of death (HR = 2.44, p = 0.0014; median OS: 9.4 vs 35.7 months). Meanwhile, for baseline QVT High patients (n=34) a decrease in QVT Score following initial treatment was associated with significantly improved survival (HR=0.30, p = 0.016; median OS: 11.5 months), relative to patients with increasing QVT Score (HR = 3.33, p = 0.016; median OS: 6.7 months).

## Discussion

Vascular complexity is a key barrier to immune cell infiltration and a determinant of therapeutic efficacy, yet no scalable clinical tools exist to quantify it. QVT Score, an automated imaging biomarker derived from routinely collected CT scans, enables both prognostication and dynamic monitoring across multiple ICI-based treatments, addressing a critical gap in precision oncology.

A wide range of strategies have been proposed for predicting ICI response using radiologic imaging.^29,30^ Traditional radiomics approaches often rely on image features developed for general computer vision tasks that are not grounded in tumor biology.^14,15^ As a result, these models frequently require indication-specific tuning and suffer from limited biological interpretability. Deep learning–based models for ICI prediction and monitoring show promise^31–34^, but they rely on opaque latent features that lack biological grounding, defy clinical interpretation,^18,35^ and often conceal spurious correlations unrelated to biological treatment response.^36^ This reality is especially troubling given the high stakes of decision-making in the cancer treatment setting.^35^ Post-hoc explainability methods offer partial output rationalization but these techniques often lack reproducibility and fail to meet clinical plausibility standards - inverting the logic of biomarker development by generating high-stakes recommendations before establishing correlative biological rationale.^18,37^

Radiogenomics attempts to improve interpretability by linking imaging to molecular biomarkers such as tumor-infiltrating CD8⁺ cells or PD-L1 status.^38–40^ While this approach enhances biological context, it ties radiomic performance to existing molecular biomarkers, constraining radiomics’ unique capacity to capture prognostic phenotypic information not accessible through conventional modalities.

In contrast, QVT Score leverages radiomic features explicitly linked to treatment-relevant biology, providing high biological interpretability and radiophenotypic characterization absent from the current biomarker landscape. Increasingly, dysregulated tumor angiogenesis has been implicated as a pivotal antagonist within the tumor microenvironment, undermining immunotherapeutic efficacy.^41^ The resulting aberrant vasculature impairs T-cell infiltration, promotes hypoxia, and drives stromal remodeling that enforces immune exclusion - collectively contributing to a tumor microenvironment hostile to antitumor immunity.^42^ Because angiogenesis shapes immune accessibility and therapeutic resistance, vascular complexity is a uniquely valuable phenotypic axis. Building on prior work demonstrating the association of individual QVT features to treatment response,^19,20,22^ our study integrates the comprehensive panel of QVT features into a unified, biologically interpretable score that characterizes NSCLC according to fundamental, treatment-agnostic phenotypes of vascular complexity. This interpretable-by-design biomarker more closely resembles molecular biomarkers such as tissue-based assays^43–45^ and circulating tumor DNA (ctDNA)^46^. However, as an automated test requiring only routinely available, non-invasive CT imaging, QVT Score offers far greater scalability and accessibility, with comparable performance to multi-modal molecular assays requiring tumor sequencing and transcriptomic profiling.^47^ In NSCLC and other cancers, immunotherapy treatment decisions remain suboptimal. PD-L1 expression, the dominant biomarker in the ICI setting, does not adequately characterize the complex dynamics of immunologic response and, consequently, poorly predicts long term survival.^48^ For example, the decision to administer mono- or combination therapy in NSCLC is driven by PD-L1 expression, with PD-L1 High (TPS>=50%) patients being de-escalated to ICI monotherapy. However, a substantial portion of these patients fail to derive durable benefit from monotherapy and could benefit from the addition of chemotherapy.^49^ Accordingly, QVT Score could aid in clinical decision-making for treatment subpopulations such as PD-L1 High, where it remained significantly associated with OS (p=0.04) and may help better identify candidates for de-escalation to ICI monotherapy.

QVT Score also offers an automated imaging-based endpoint for real-time treatment efficacy monitoring. Standard response assessments such as RECIST correlate poorly with OS in the immunotherapy setting, largely due to atypical response patterns, including pseudoprogression. Early t on-treatment changes in QVT Score were significantly associated with long-term survival outcomes, measuring vascular remodeling reflective of biological response to treatment. Importantly, these associations were independent of RECIST-defined best overall response categories and volumetric tumor burden changes,^50^ highlighting the unique biological information captured by QVT Score. This enables faster clinical decision-making and treatment adaptation, such as timely escalation to combination therapy in patients unlikely to benefit from monotherapy.

In drug development, QVT Score can serve as both a non-invasive baseline risk-screening tool and a surrogate endpoint for early treatment efficacy, supporting adaptive trial designs across diverse drug classes. Our cloud-based pipeline delivers a fully automated QVT Score in an average of less than 30 minutes from clinical imaging. For agents targeting vascular biology, such as VEGF inhibitors or VEGF-ICI combinations, QVT Score offers a precise, interpretable endpoint to quantify mechanism-of-action and track response. We observed that in patients with elevated vascularity at baseline, treatment-induced reduction in QVT Score were linked to improved outcomes. Our findings highlight that successful early vascular normalization can mitigate the adverse prognosis of unchecked tumor angiogenesis and is detectable through routine clinical imaging.

This study did have its limitations. First, due to the availability of longitudinal data, it was necessary to pool patients from both the discovery and validation datasets to form a longitudinal evaluation cohort. While evaluating the score as a monitoring tool in an independent validation cohort would be more robust, QVT Score is less susceptible to overfitting than traditional machine learning models due to its unsupervised nature and the use of only pretreatment scans for development. Second, this study employed retrospective, real world data for development and validation of the QVT Score. Substantial heterogeneity existed across imaging protocols, follow-up duration, and clinical demographics. While image pre-processing and normalization strategies accounted for this variation, further validation to control for these factors, such as in randomized clinical trials (RCTs), is warranted and ongoing.

Future directions include extending vascular phenotyping to other tumor types, validating clinical utility and pursuing regulatory qualification of the QVT Score for clinical workflow integration. Given its scalability, interpretability, and mechanistic grounding, QVT Score can enable precision treatment, guide angiogenesis-targeted therapy and accelerate oncology drug development.

## Data Availability

The private datasets analyzed in this study are not publicly available due to restrictions imposed by data-sharing agreements with the contributing institutions. Access to these datasets is therefore not possible at this time.

## Ethics approval and consent to participate

This study was reviewed and approved by the Institutional Review Board, which determined that it met the criteria for exemption under 45 CFR 46.104(d)(4). The research involved the secondary use of existing clinical and imaging data, recorded in such a manner that subjects could not be identified directly or through linked identifiers. No new data were collected, and no contact or intervention with participants occurred.

## Acknowledgements

We extend our deepest gratitude to the patients who contributed data towards this project. We also thank the National Institutes of Health (NIH), the United States Department of Veterans Affairs (VA), the National Science Foundation (NSF), and the Department of Defense (DOD) for funding the foundational academic research preceding this study.

## Supplementary Materials

**Supplementary Figure 1.**
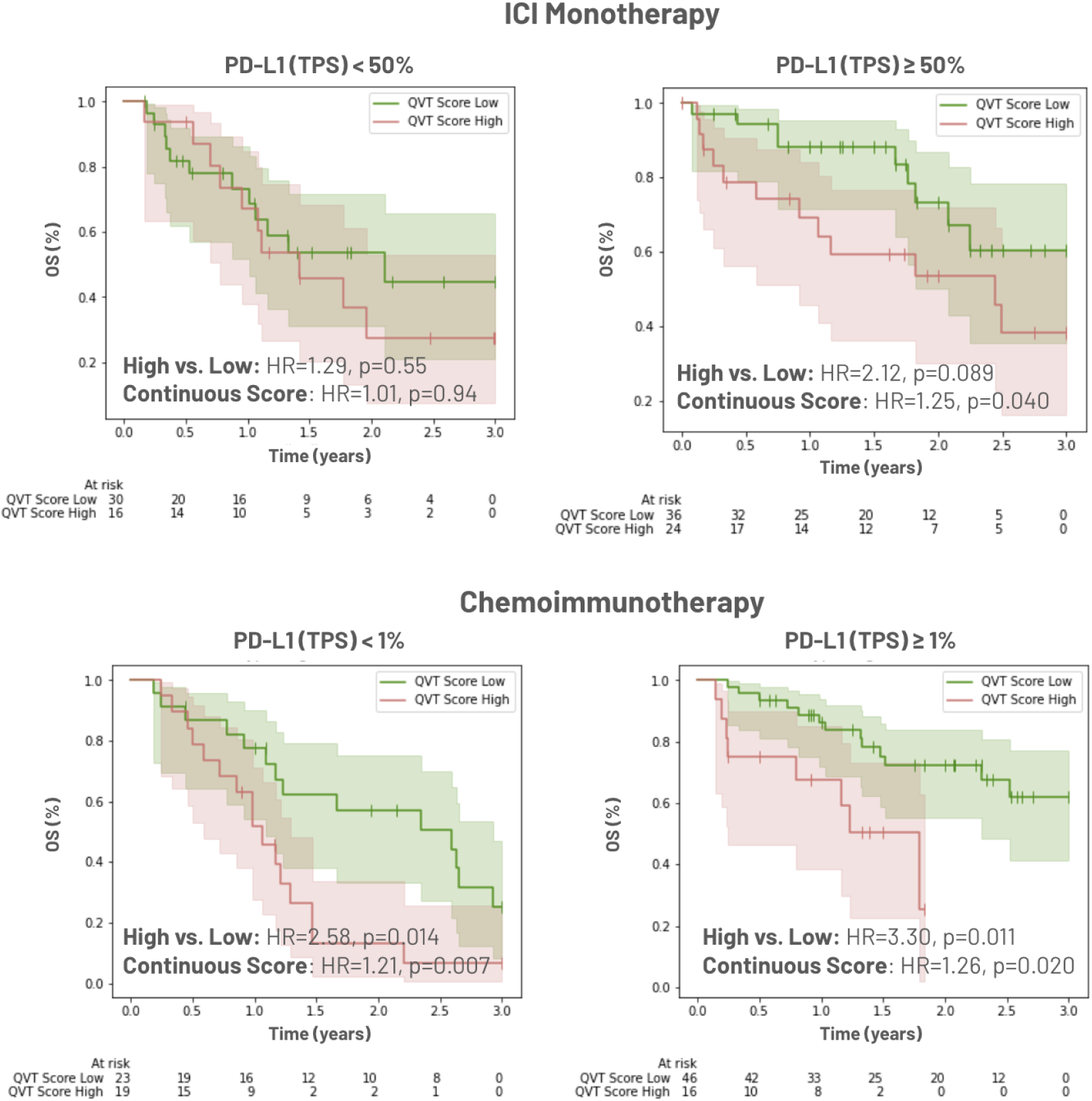
Kaplan Meier curves depicting OS for QVT Score groups within PD-L1 expression subsets. For each treatment, PD-L1 threshold was chosen according to the most established cutoff for that indication.

